# The impact of diabetes mellitus and HBV infection on major adverse cardiovascular events related to statin use for Chinese population with cardiovascular disease

**DOI:** 10.1101/2024.11.05.24316797

**Authors:** Qian Wu, Xiaoxuan He, Xiaoning Tong, Ying Li, Xiaoqin Wang

**Author notes:** He contributed equally to this work (Qian Wu, Xiaoxuan He). **Correspondence to:** Dr. Xiaoqin Wang, M.D. Department of Clinical Laboratory, The First Affiliated Hospital of Xi’an Jiaotong University, No. 277 West Yanta Road, Xi’an 710061, Shaanxi Province, China.

## Abstract

**BACKGROUND:** The association between hepatitis B virus (HBV) infection, diabetes mellitus (DM) and cardiovascular disease (CVD) remains uncertain. This study was aimed to investigate the impact of HBV infection and DM on 13 major adverse cardiovascular events (MACEs) among CVD patients with or without statin use.

**METHODS AND RESULTS:** In total, 45,013 individuals participated in the baseline survey between June 2020 and August 2023. The patients were categorized into two groups according to their surface antigen status. Finally, a sample size of 496 participants was included in the study: patients with coexisting CVD and HBV (n = 225) and patients with CVD alone (n = 271).The implementation of statins has demonstrated the potential to reduce the incidence of coronary heart disease (CHD) and angina pectoris (AP) in individuals with CVD who are also affected by HBV infection. The analysis of stratification, taking into account the presence of DM, has demonstrated a significant increased vulnerability to myocardial infarction (MI), acute myocardial infarction (AMI), and heart failure (HF) among individuals with HBV infection combined with CVD. DM and hemoglobin A1c (HBA1c) level of ≥ 6 are identified as risk factors for the development of CHD, MI, AP, and hypertension. Our study also suggests that statin therapy led to a dose-dependent decrease in liver fibrosis, as well as improvement of the atherogenicity index and systemic inflammatory response in patients with CVD.

**CONCLUSION:** Our study indicates that CVD patients who also have HBV infection or DM may experience a reduction in MACEs when treated with statins. Furthermore, our research indicates that focusing on long-term glycemic control is essential in this patient population in order to enhance cardiovascular outcomes. The observed improvements in hepatic function, atherosclerotic burden, and systemic inflammation associated with statin therapy may contribute to the favorable cardiovascular outcomes among individuals with CVD.

**Clinical Perspective:** *What Is New?:*

- This study investigates the association between statin therapy and major adverse cardiovascular events (MACEs) in Chinese patients with cardiovascular disease who have concurrent hepatitis B virus infection or diabetes mellitus.
- The findings suggest potential cardiovascular benefits of statin therapy in this specific patient population.
- The study explores the relationship between statin use and changes in hepatic function, atherosclerotic markers, and inflammatory parameters.

*What Are the Clinical Implications?:*

- The results provide evidence supporting statin use in cardiovascular disease patients with hepatitis B virus infection or diabetes mellitus.
- Glycemic control appears to be an important factor in optimizing cardiovascular outcomes in these patients.
- Regular monitoring of hepatic and inflammatory markers may be valuable in assessing treatment response.

---

**C**oronary heart disease (CHD) is the primary cause of mortality in humans, accounting for approximately 9 million deaths annually worldwide ^1^. CHD encompasses cardiovascular conditions resulting from the narrowing or obstruction of the coronary arteries due to atherosclerosis, as well as myocardial ischemia, hypoxia, or necrosis caused by functional alterations in these arteries ^2^. Presently, there exists a prevailing belief within the academic community that the presence of obesity, smoking, and alcohol consumption plays a substantial role in the advancement of CHD ^3, 4^. Furthermore, it is worth emphasizing that hypertension, hyperlipidemia, hypercholesterolemia, and diabetes mellitus (DM) also contribute to the development of CHD ^5–8^. Notably, DM represents a significant risk factor, as the elevated blood glucose levels experienced by individuals with this condition often induce oxidative stress and inflammatory harm, ultimately resulting in atherosclerosis, coronary artery dysfunction, myocardial infarction, and cellular impairment, thereby fostering the progression of CHD ^9^. DM is additionally linked to an unfavorable long-term prognosis in individuals diagnosed with CHD. In comparison to patients without DM, those with DM complicated by CHD exhibit a higher mortality rate over a span of 10 years ^10^.

The global prevalence of Hepatitis B virus (HBV) infection is substantial, rendering it a significant concern for public health. Prolonged HBV infection can result in chronic hepatitis, cirrhosis, liver decompensation, and hepatocellular carcinoma ^11^. Chronic HBV infection is characterized by a state of inflammation, which has been observed to elevate the likelihood of major adverse cardiovascular events (MACE) and cerebrovascular disease in other conditions featuring chronic low-grade inflammation ^12, 13^. Several studies have examined the association between HBV infection and cardiovascular disease (CVD); however, the findings have been inconclusive ^14–17^. Certain studies have identified a positive correlation between HBV infection and coronary artery disease ^17^, whereas others have indicated a negative correlation ^14–16^. In individuals diagnosed with chronic viral hepatitis, it has been observed that those with HCV infection exhibit a notably elevated susceptibility to composite arterial events and all-cause mortality when compared to those with HBV infection ^18, 19^. Furthermore, investigations have indicated that there is no substantial association between the prevalence of macrovascular complications, microvascular complications of diabetes, diabetic ketosis, or diabetic ketoacidosis and the status of HBV infection ^20^. Hence, it is crucial to persist in the examination of the correlation between HBV infection and MACE.

Statins have demonstrated efficacy in mitigating the occurrence of CVD and are endorsed as the primary therapeutic approach for hypercholesterolemia in clinical guidelines ^21, 22^. The American College of Cardiology/American Heart Association guidelines for managing blood cholesterol levels to minimize the risk of atherosclerotic CVD (ASCVD) in adults have classified statins into low, moderate, and high potency categories, based on comprehensive analysis of their lipid-lowering effects across various clinical trials and direct comparisons. High-potency statins, such as rosuvastatin at doses of 20 mg/d and 40 mg/d, and atorvastatin at doses of 40 mg/d and 80 mg/d, have exhibited a noteworthy decrease in low-density lipoprotein cholesterol (LDL-C) levels surpassing 50% ^21^. Additionally, decreased cholesterol levels by statin use may impact the advancement of hepatic fibrosis by ameliorating portal hypertension and reducing the excessive accumulation of hepatic triglycerides ^23–25^. Numerous clinical studies have demonstrated the efficacy of statin administration in patients with NAFLD, leading to improvements in liver ultrasonography and liver function tests ^26–28^. The independent association of statin use with reductions in hepatic steatosis and fibrosis stage in NAFLD patients has also been observed ^26, 28^. The pathogenesis of hepatic fibrosis is characterized by a multifaceted interaction between lipid metabolism and systemic chronic inflammatory response in patients with CVD ^29, 30^.

Hence, this study aimed to gather data on patients with both HBV and CVD, as well as those with CVD alone, in order to examine the correlation between statin use and HBV infection, DM, or hemoglobin A1c (HbA1c) levels among individuals with CVD. Additionally, the study sought to assess the impact of HBV infection, DM, HbA1c levels, or statin use on various cardiovascular outcomes in patients with CVD. Currently, there is uncertainty regarding the impact of statins on hepatic fibrosis and systemic inflammatory response, as well as their effects on CVD and lipid metabolism. The identification of specific markers in this context remains ambiguous. It is our assertion that the identification of suitable markers is crucial for the monitoring of liver fibrosis and systemic inflammation in patients with CVD undergoing statin therapy. Our research aimed to assess the impact of statin therapy on hepatic fibrosis, systemic inflammation, and atherogenic indices in patients with CVD.

## MATERIALS AND METHODS

### Study design and participants

A prospective cohort study was conducted to examine the prevalence, incidence, and associated factors, such as HBV infection, in relation to CVD. The study was approved by the Ethics Committee of the First Affiliated Hospital of Xi’an Jiaotong University (No:XJTU1AF2024LSYY-054). In total, 45,013 individuals participated in the baseline survey at the First Affiliated Hospital of Xi’an Jiaotong University (Xi’an, China) between June 2020 and August 2023. Of these, 43,750 cases were excluded due to HCV, HIV, syphilis infection, as well as incomplete basic data of test results and medical records. A total of 268 patients presenting with hepatic hemangioma, toxic diffuse goiter with hyperthyroidism, hypothyroidism, endometrial cancer, stage 5 hemodialysis status of chronic kidney disease, chronic renal insufficiency, skull osteoma, and right lobe cyst of thyroid gland were excluded from the study. Additionally, 499 cases with incomplete test data, including five quantitative variables (disease load, blood cells, blood lipid, and other information), as well as incomplete drug use information, were also excluded. Consequently, a final sample size of 496 participants was included in the study (refer to Figure 1). The patients were categorized into two groups according to their surface antigen status: patients with coexisting CVD and HBV (n = 225) and patients with CVD alone (n = 271). In addition, a sample size of 494 CVD participants was included, patients with statin use (n = 415) and statin-free (n = 79), to investigate the effect of statin use on hepatic fibrosis, atherosclerotic burden, and systemic inflammation.

**Figure 1.**
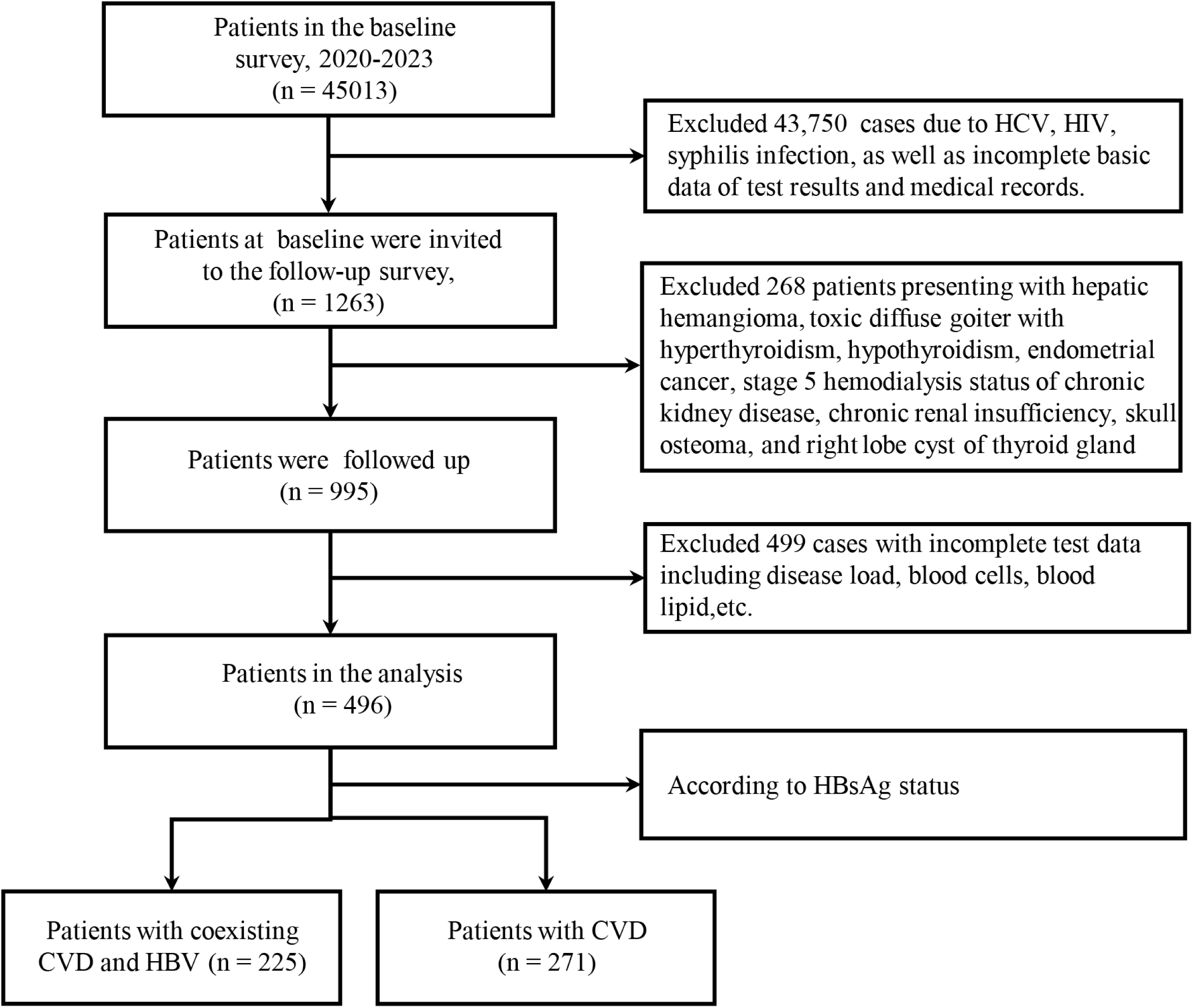
Flow diagram of cohort selection. Consequently, a final sample size of 496 participants was included in the study. The patients were categorized into two groups according to their surface antigen status: patients with coexisting CVD and HBV (n = 225) and patients with CVD alone (n = 271). CVD, cardiovascular disease; HBV, hepatitis B virus; HBsAg, hepatitis B surface antigen.

### Clinical data collection and measurements

The variables considered in this study encompassed patient demographics such as age and gender, as well as medical factors including hypertension, smoking/alcohol consumption, diabetes, disease status, biochemical indicators, blood cell indicators, HBV five-item quantitative, HBV-DNA quantitative, statin use, antiviral therapy for hepatitis B, and major adverse cardiovascular events. The ratios of platelet count to lymphocyte count (PLR) and neutrophil count to lymphocyte count (NLR) are computed to assess certain adverse cardiovascular events, such as recurrent angina pectoris (AP), hospitalization for unstable angina pectoris (UAP), acute myocardial infarction (AMI), severe arrhythmia, heart failure (HF), and death due to CHD. Major adverse cardiovascular events include recurrent AP, UAP requiring hospitalization, AMI, severe arrhythmia, HF, coronary-related death, etc. Secondary adverse cardiovascular events include Killip classification Ⅱ-Ⅳ or NYHA Classification Ⅱ-Ⅳ.

All specimens were tested within 2 hours following their collection. In cases where immediate testing was not feasible, specimens were stored at a temperature of -20°C for a duration not exceeding 2 days. The fluorescence quantitative PCR detection of HBV-DNA load was conducted utilizing the 7500 fluorescence quantitative PCR instrument and associated reagents (ABI, USA). Quantitative determination of biochemical indicators was performed using the Roche AU5600 automatic biochemical analyzer and its corresponding original reagents. The quantification of the five components of serum hepatitis B was performed using the i2000sr instrument manufactured by Abbott Laboratories, USA. The detection of blood cell indicators was conducted using the routine automatic flow cytometer Mindray CAL8000. The Roche E801 automatic immunoassay analyzer, along with original reagents, was utilized for the quantitative determination of B-type natriuretic peptide (BNP) in the anterior brain. HbA1c was assessed with high-performance liquid chromatography. All aforementioned measurements were performed in accordance with the manufacturer’s instructions.

### Calculations of the albumin-bilirubin index (ALBI) score, fibrosis 4 index (FIB-4) index, complete blood count derived inflammation index (CBCIIs) and atherogenic indices

Non-invasive and non-destructive testing markers, including FIB-4, ALBI, and CBCIIs, such as platelet monocyte ratio (PMR), neutrophil/lymphocyte ratio (NLR), SII, systemic inflammatory response index (SIRI), and aggregate index of systemic inflammation, and atherogenic indices, such as atherogenic index of plasma (AIP), atherogenic index (AI), Castelli risk index-I (CRI-I), Castelli risk index-II (CRI-II), and triglyceride-rich lipoprotein cholesterol (TRL-C) were used to evaluate the effects of statin use on hepatic fibrosis, systemic inflammation, and arteriosclerotic degree in patients with CVD. The calculation formulas of hepatic fibrosis indicators ALBI score, FIB-4 index were measured as described previously ^31^. CBCIIs, including SIRI, SII, aggregate index of systemic inflammation (AISI), SIRI/HDL-C, and SIRI×LDL-C, were calculated as described previously ^32, 33^. Atherogenic indices, such as adjusted AIP, AIP, AI, CRI-I, CRI-II, LCI, and TRL-C, were calculated as described previously ^34–37^.

### Statistical analysis

In the case of measurement data that conforms to a normal distribution, the mean ± standard deviation is employed for representation, and intergroup comparison is carried out through the utilization of a t-test. Conversely, for measurement data that does not adhere to a normal distribution, the median (quartile) [M (P_25_ ∼ P_75_)] is employed for representation, and intergroup comparison is conducted using the Mann-Whitney U non-parametric test. Count data is represented using frequency and percentage (%), and intergroup comparison is performed using either the chi-square test or Fisher’s exact probability method. The Breslow-Day test is employed for conformance testing in hierarchical analysis, while the Mantel-Haenszel test is utilized for hierarchical chi-square testing. To address potential confounding, logistic regression models were used to estimate the odds ratio (OR) and 95% CI for MACE. All statistical analyses were conducted using SPSS, version 22.0 (SPSS Inc, USA). A 2-tailed *P* value of less than 0.05 was considered to be statistically significant.

## RESULTS

**Table 1** presents the demographic characteristics of the study participants. A total of 496 subjects were included in the analysis, comprising 225 individuals with pure CVD and 271 individuals with CVD accompanied by HBV infection. Notably, the CVD combined with HBV infection group exhibited a statistically significant increase in the proportion of males and AMI when compared to the CVD alone group (both P < 0.05). Furthermore, when comparing patients with CVD alone to those with CVD combined with HBV, it was observed that the latter group exhibited elevated levels of HBsAg, HbsAb, and HbcAb (all *P* < 0.05). Conversely, the CVD combined with HBV group displayed lower levels of weight, medication dosage, TG, Lpa, PLR, and PLT in comparison to the CVD alone group. Additionally, the proportions of diabetes and pre-hospital medication were relatively lower in the CVD combined with HBV group. Notably, the remaining indicators presented in **Table 1** did not demonstrate statistical significance. The quantitative (**Supplementary table 1**) and qualitative (**Supplementary table 2**) baseline characteristics and statistical comparisons of individuals with CVD who were either using or not using statins were also documented. It is evident that there are statistically significant differences in height, weight, platelet count, alanine aminotransferase levels, and aspartate aminotransferase levels among the three groups (all *P* < 0.05). The utilization of statins resulted in lower TG (*P* = 0.000), LDL-C (*P* = 0.000), ApoB (*P* = 0.000), and non-HDL-C (*P* = 0.000) levels. Based on the findings presented in **Supplementary table 2**, it is evident that there exist statistically significant disparities in gender, diabetes, and smoking history across the three groups, with or without statin usage (all *P* < 0.05). It is worth noting that long-term use of statins has been associated with an increased risk of DM ^38^. Consequently, clinicians may be avoiding prescribing statins to cardiovascular patients with diabetes, leading to a higher proportion of non-statin use in this population (65.8%). Conversely, no statistically significant variations were observed in hypertension, alcohol history, and anti-HBV treatment among the three groups (all *P* > 0.05).

**Table 1.**
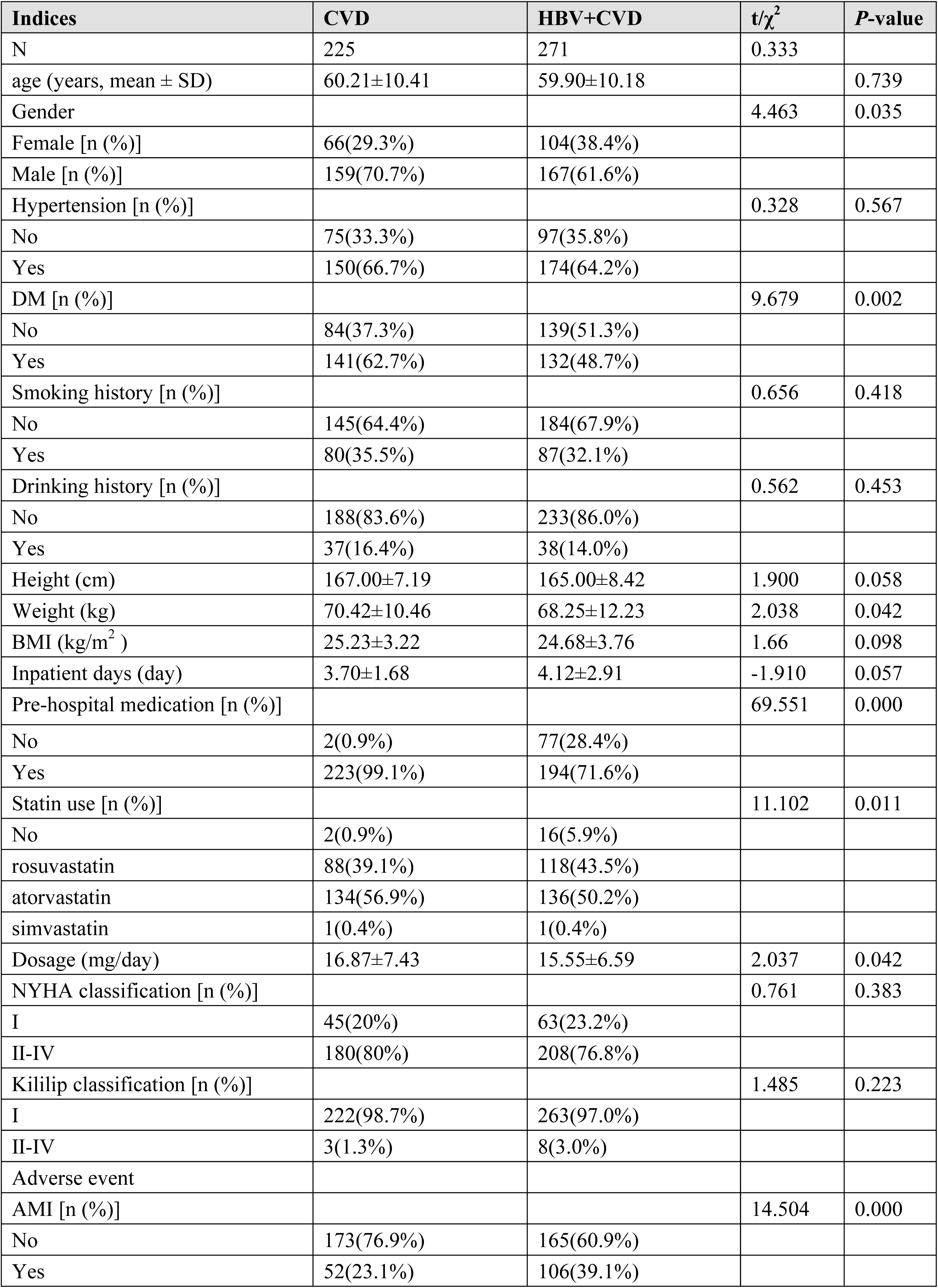
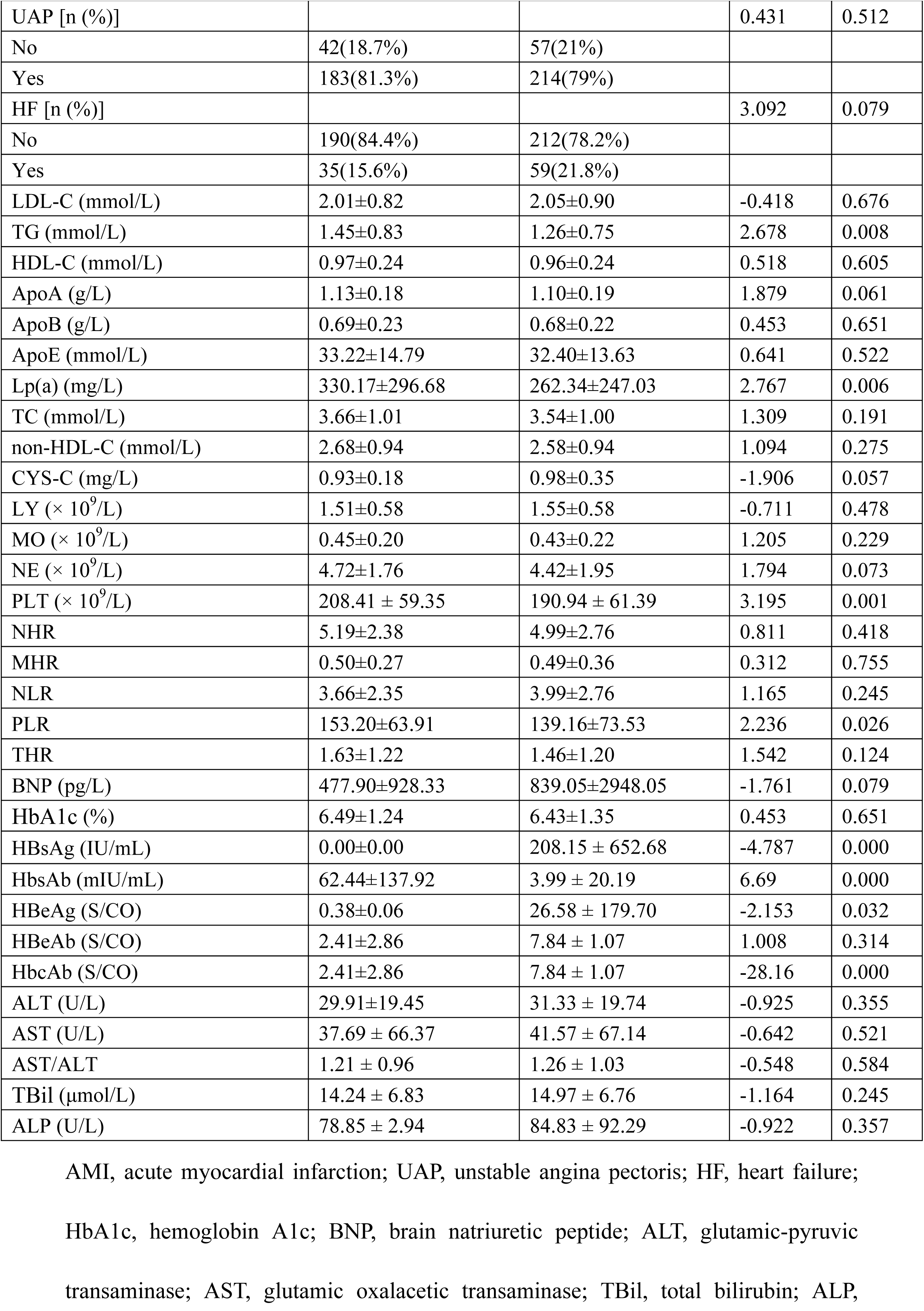
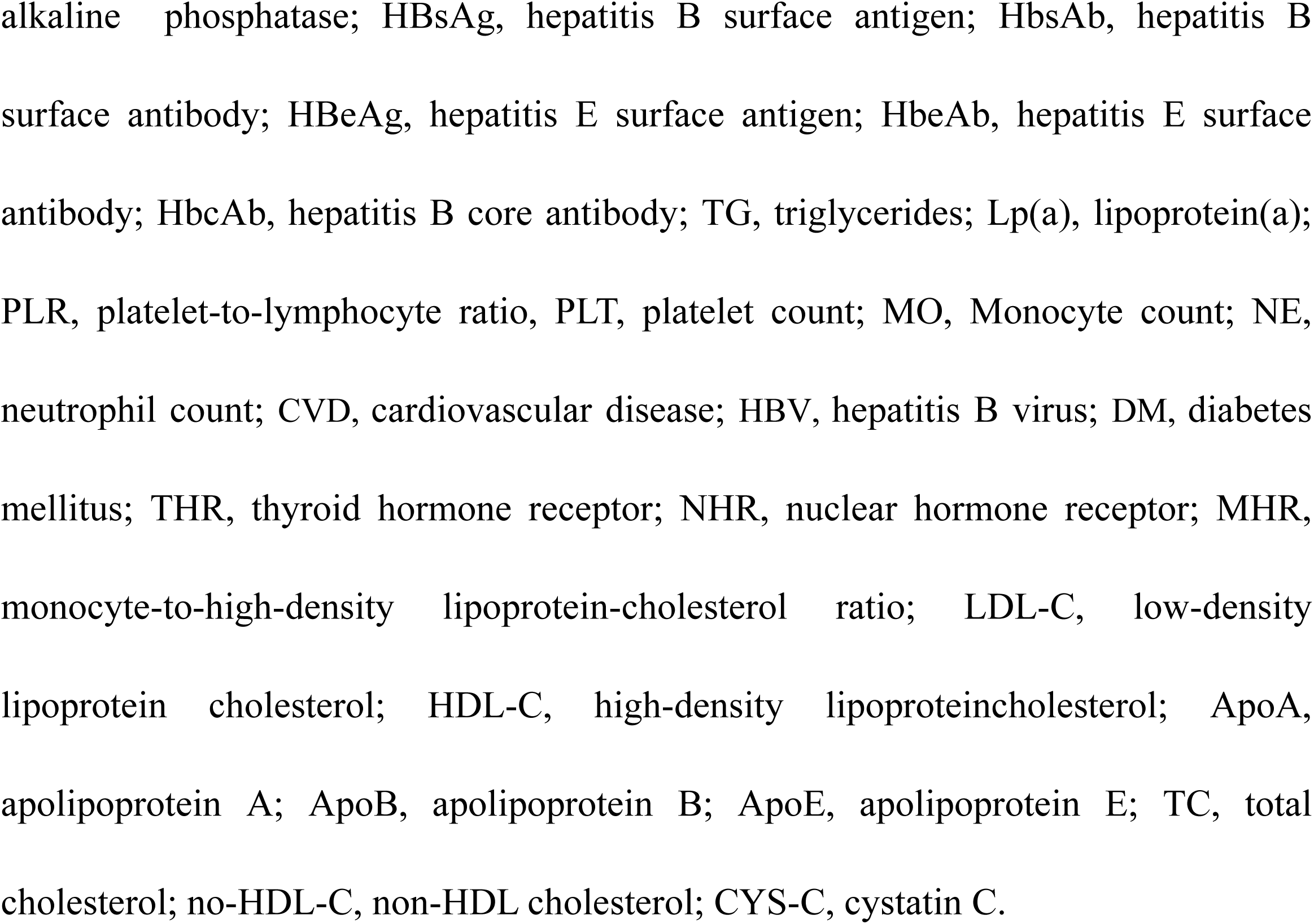
Baseline characteristics of CVD with HBV and CVD alone participants.

### Relationship of Statin Use with HBV Infection and Quantification

To examine the relationship between statin use and HBV infection, we conducted a study comparing HBV infection rates and quantitative parameters in patients both with and without statin use. Our analysis revealed a significant association between statin use and HBV infection rates (χ^2^ = 76.835, *P* = 0.000). Subsequent pairwise comparisons demonstrated that the HBV infection rate in the group not using statins was 97.5% (77/79), whereas the rate in the group using atorvastatin 20 mg was 40.4% (91/225), indicating a protective effect with an OR of 0.018 (95% CI: 0.004 - 0.074). In the rosuvastatin 10 mg group, the HBV infection rate was 53.7% (102/190), indicating a potential protective effect with an OR of 0.030 (95% CI: 0.007 - 0.126). Furthermore, the HBV infection rate was 40.4% (91/225) in the atorvastatin 20 mg group and 53.7% (102/190) in the rosuvastatin 10 mg group, demonstrating a statistically significant difference between the two groups (*P* = 0.002). This suggests that the HBV infection rate in the rosuvastatin 10mg group was relatively higher, with an odds ratio of 1.94 (95% CI: 1.28 - 2.94) (**Table 2**). The HbsAg levels ranged from less than 0.05 to 250, the Median Difference (0-1) was calculated to be 70 (95% CI: 26 to 125). Additionally, the Median Difference (0-2) was found to be 63 (95% CI: 18 to 117). These findings indicate that the quantification of HbsAg in individuals using statins is notably reduced compared to those not using statins (**Table 3**). The analysis of the Median Difference (1-2) yielded a value of 0 (95% CI: 0 to 0), suggesting a lack of statistically significant disparity in the effectiveness of the two statins in reducing HbsAg quantitation. However, the OR (atorvastatin/rosuvastatin) in Table 2 was calculated as 1.94 (95% CI: 1.28 to 2.94), indicating a lower infection rate associated with atorvastatin (**Table 3**). This apparent contradiction underscores the need for a nuanced interpretation that considers both qualitative and quantitative aspects of the investigation. Furthermore, when compared to the group not using statins, the quantification of HBV-DNA and HBcAb was significantly lower in the statins use group. Conversely, the quantification of HBsAb, HBeAg, and HBeAb was significantly higher in the statins use group (**Table 3**). In addition, **Table 4** demonstrates a significant dose-effect relationship between statin dosage and HbsAg quantification (*P* = 0.000), indicating that higher doses result in lower HbsAg levels, with the exception of a slight rebound observed at the highest dose of rosuvastatin 20 mg. Nevertheless, the limited sample size at this dosage level necessitates further validation of these findings.

**Table 2.** Relationship between statin use and HBV infection.

| Statins | HBV infection | | Total (%) | $\chi^2$ | P-value<br>(two-sided) |
| --- | --- | --- | --- | --- | --- |
|  | negative | positive |  |  |  |
| no use (pre-admission) | 2 (2.5%) | 77 (97.5%) | 79 (100%) | 76.835 | 0.000 |
| atorvastatin | 134 (59.6%) | 91 (40.4%) | 225 (100%) |  |  |
| rosuvastatin | 88 (46.3%) | 102 (53.7%) | 190 (100%) |  |  |
| total | 224 | 270 | 494 |  |  |
| OR (no use/atorvastatin) | 0.018 (95% CI: 0.004 to 0.074) |  |  | 76.904 | 0.000 |
| OR (no use/rosuvastatin) | 0.030 (95% CI: 0.007 to 0.126) |  |  | 48.048 | 0.000 |
| OR (atorvastatin/rosuvastatin) | 1.940 (95% CI: 1.280 to 2.940) |  |  | 9.979 | 0.002 |

**Table 3.** Relationship between statin use and quantitative parameters of HBV.

| Variables | Statins use | N | P50 (P25 – P75) | Mean ± SD | Range |
| --- | --- | --- | --- | --- | --- |
| HBsAg | No use=0 | 79 | >250 (26 to >250) |  | <0.05 to >250 |
|  | atorvastatin =1 | 225 | <0.05(<0.05 to 129) |  | <0.05 to >250 |
|  | rosuvastatin=2 | 190 | 2(<0.05 to 174) |  | <0.05 to >250 |
|  | measure |  | H = 65.606 |  |  |
|  | P |  | 0.000 |  |  |
| Median Difference(0-1) |  |  | 70(95% CI: 26 to 125) |  |  |
| Median Difference(0-2) |  |  | 63(95% CI: 18 to 117) |  |  |
| Median Difference(1-2) |  |  | 0(95% CI: 0 to 0) |  |  |
| HBsAb | No use=0 | 73 | 0.58(0.20 to 1.45) | 2.4±7.9 | 0 to 52 |
|  | atorvastatin =1 | 199 | 2.10(0.58 to 20.97) | 48±133 | 0 to 951 |
|  | rosuvastatin=2 | 181 | 1.31(0.43 to 5.98) | 21±57 | 0 to 338 |
|  | measure |  | H = 31.257 |  |  |
|  | P |  | 0.000 |  |  |
| HBeAg | No use=0 | 73 | 0.34(0.31 to 0.42) | 0.68±2.18 | 0.01 to 19 |
|  | atorvastatin =1 | 214 | 0.39(0.34 to 0.43) | 9±111 | 0.01 to 1617 |
|  | rosuvastatin=2 | 186 | 0.39(0.34 to 0.43) | 26±175 | 0.0225 to 1438 |
|  | measure |  | H = 8.044 |  |  |
|  | P |  | 0.018 |  |  |
| HBeAb | No use=0 | 73 | 0.01(0.01 to 0.02) | 0.20±0.48 | 0.01 to 2 |
|  | atorvastatin =1 | 214 | 0.86(0.02 to 1.76) | 1.17±3.07 | 0 to 43 |
|  | rosuvastatin=2 | 186 | 0.47(0.01 to 1.74) | 1.75±6.27 | 0.01 to 51 |
|  | measure |  | H = 47.239 |  |  |
|  | P |  | 0.000 |  |  |
| HBcAb | No use=0 | 73 | 7.89(7.09 to 8.61) | 7.69±1.57 | 0.06 to 10 |
|  | atorvastatin =1 | 214 | 6.06(0.16 to 7.62) | 4.65±3.46 | 0.04 to 11 |
|  | rosuvastatin=2 | 186 | 7.02(0.62 to 8.04) | 5.26±3.53 | 0.03 to 11 |
|  | measure |  | H = 47.295 |  |  |
|  | P |  | 0.000 |  |  |
| HBV DNA | No use=0 | 42 | 129(<100 to 2103) |  | <10 to 11800000 |
|  | atorvastatin =1 | 121 | <100(<100 to <100) |  | <10 to 1310000 |
|  | rosuvastatin=2 | 111 | <100(<100 to <100) |  | <10 to 476000000 |
|  | measure |  | H = 5.522 |  |  |
|  | P |  | 0.063 |  |  |

**Table 4.** Relationship between statin dose and HbsAg quantification.

| Statins use | HbsAg quantification |  |  | J-T | P (two-sided) |
| --- | --- | --- | --- | --- | --- |
|  | N | P50(P25–P75) | Range |  |  |
| no use (pre-admission) | 79 | 251(26 to >250) | <0.05 to >250 |  |  |
| atorvastatin (10 mg) | 19 | 78(<0.05 to >250) | <0.05 to >250 |  |  |
| atorvastatin (20 mg) | 182 | <0.05 (<0.05 to 83) | <0.05 to >250 |  |  |
| atorvastatin (40 mg) | 19 | <0.05 (<0.05 to >250) | <0.05 to >250 | -7.615 | 0.000 |
| rosuvastatin (10 mg) | 8 | 17(<0.05 to 198) | <0.05 to >250 |  |  |
| rosuvastatin (20 mg) | 166 | 1(<0.05 to 112) | <0.05 to >250 |  |  |
| rosuvastatin (40 mg) | 12 | 80(<0.05 to >250) | <0.05 to >250 | -5.936 | 0.000 |

### The impact of HBV infection, stratified by statins use, on multiple clinical outcomes among CVD patients

In our study, atorvastatin and rosuvastatin emerge as the prevailing lipid-lowering medications, with atorvastatin 20 mg and rosuvastatin 10 mg serving as the primary drug dosage combinations. This particular combination is employed as a stratification factor to examine the impact of HBV infection on 13 clinical outcomes among individuals with CVD. According to the findings depicted in **Figure 2**, it was observed that HBV infection exhibited a protective effect on the occurrence of CHD (OR = 0.27, 95% CI: 0.12 – 0.60; *P* = 0.001) and AP (OR = 0.56, 95% CI: 0.36 – 0.86; *P* = 0.008). Conversely, HBV infection was identified as a risk factor for AMI (OR = 2.24, 95% CI: 1.40 – 3.57; *P* = 0.001). Furthermore, it was determined that HBV infection did not significantly impact the remaining 11 clinical outcomes in individuals with CAD.

**Figure 2.**
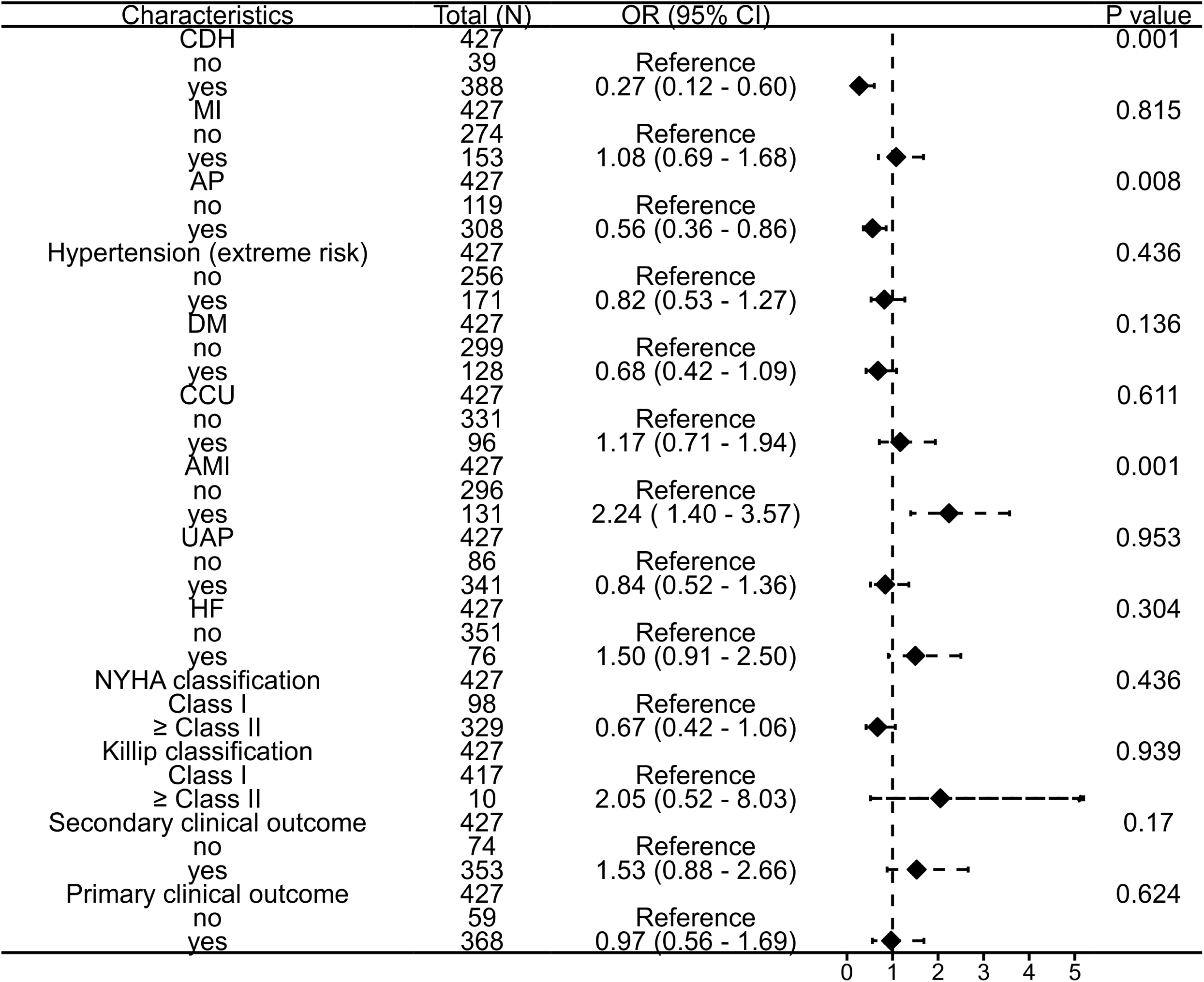
The impact of HBV infection, stratified by statins use, on multiple clinical outcomes among CVD patients. OR, odds ratio; CDH, coronary heart disease; MI, myocardial infarction; AP, angina pectoris; DM, diabetes mellitus; CCU, coronary care unit; AMI, acute myocardial infarction; UAP, unstable angina pectoris; HF, heart failure.

### The impact of HBV infection, stratified by DM, on multiple clinical outcomes among CVD patients

Given that the prevalence of DM among patients with and without HBV infection was 48.7% and 62.7%, respectively (*P* = 0.002; **Table 1**), it is evident that the baseline characteristics were not equivalent. Consequently, the presence of DM may exert an influence on cardiovascular outcomes. In this study, DM was employed as a stratified variable to examine the potential impact of HBV infection on clinical outcomes in patients with CVD. The results of our study indicate that the coexistence of DM and HBV infection may significantly increase the risk of AMI (OR = 5.37, 95% CI: 2.91 - 9.92; *P* < 0.001) as well as unfavorable secondary clinical outcomes (OR = 4.34, 95% CI: 2.06 - 9.15; *P* < 0.001) (**Figure 3**). These findings suggest the importance of implementing strict blood sugar control and anti-HBV therapy in patients with this comorbidity.

**Figure 3.**
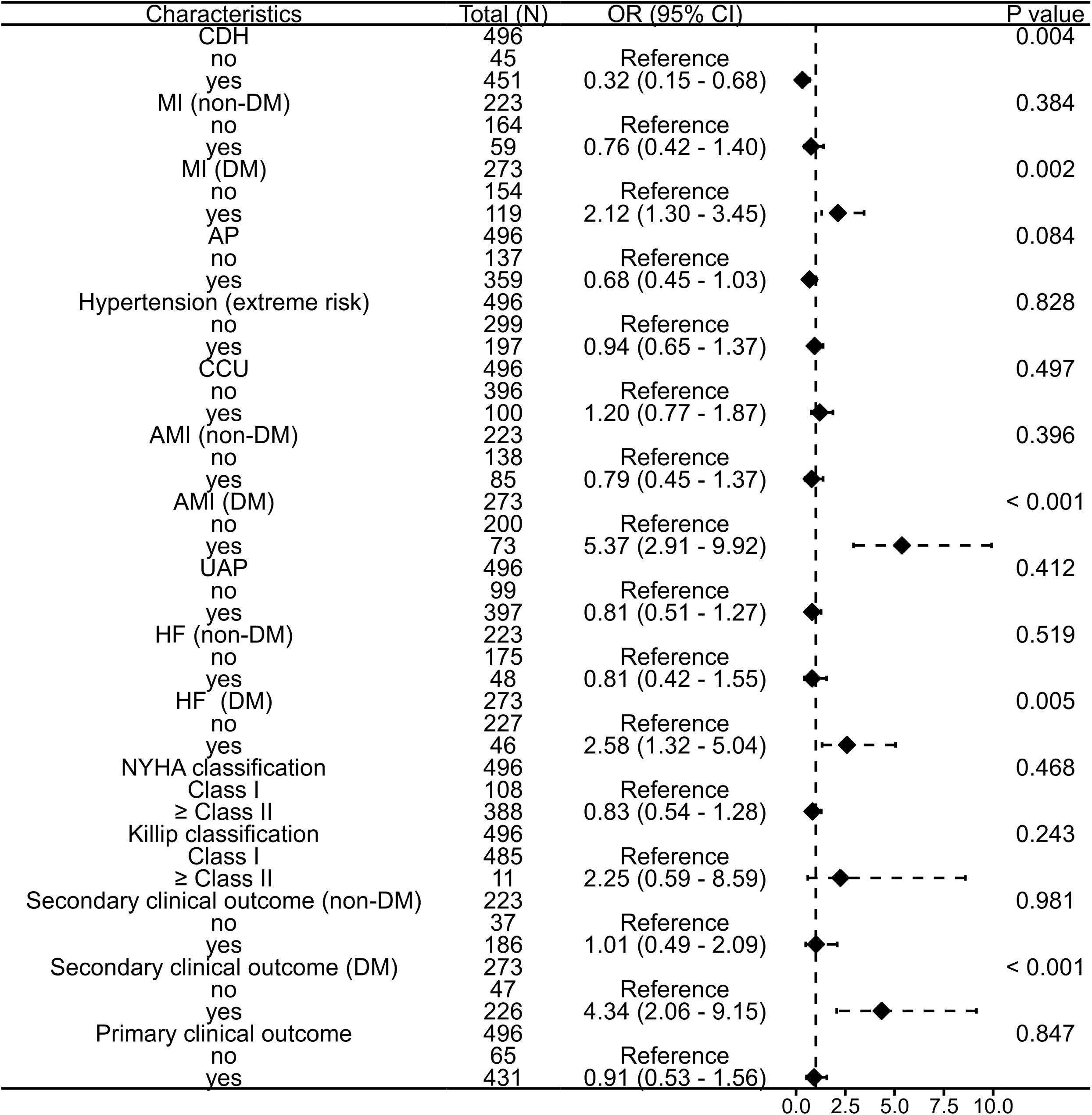
The impact of HBV infection, stratified by DM, on multiple clinical outcomes among CVD patients. OR, odds ratio; CDH, coronary heart disease; MI, myocardial infarction; AP, angina pectoris; DM, diabetes mellitus; CCU, coronary care unit; AMI, acute myocardial infarction; UAP, unstable angina pectoris; HF, heart failure.

### The impact of DM and HbA1c (≥ 6% or < 6%) on multiple clinical outcomes among CVD patients

Based on the baseline characteristics of the population statistics, a comprehensive analysis was conducted on a total of 496 individuals, comprising 273 subjects with DM and 223 subjects without DM. The findings revealed that, in comparison to the non-DM group, the DM group exhibited a notably higher prevalence of hypertension history and the administration of atorvastatin 20 mg dosage (both *P* < 0.05). Conversely, the prevalence of smoking history, rosuvastatin 10 mg dosage, and overall statin usage were significantly lower in the DM group (all *P* < 0.05). Furthermore, in comparison to the non-DM group, the DM group exhibited elevated levels of MO, NE, MHR, NHR, BNP, ALS, and HbA1c (all *P* < 0.05). Conversely, the DM group displayed a relatively diminished ApoE level when contrasted with the non-DM group. There were no statistically significant differences observed in various demographic and clinical factors, including age, gender, height, weight, BMI, alcohol consumption history, LDL-C level, TG level, HDL-C level, triglyceride-to-HDL-C ratio, ApoA level, ApoB level, Lp(a) level, TC level, non-HDL-C level, CYS-C level, LY count, PLT count, NLR, PLR, AST/ALT, AST level, TBil level, and ALP level, between the two groups (**Supplementary table 3 and 4**). As shown in **Figure 4**, DM poses a significant risk for CHD (RO = 6.9, 95% CI: 3.2-14.9; *P* < 0.001) among patients with CVD, MI (RO = 2.2, 95% CI: 1.5-3.2; *P* < 0.001), AP patients who were not prescribed statins prior to admission (RO = 3.5, 95% CI: 1.3-9.3; *P* = 0.01), and individuals with hypertension extreme risk (RO = 2.9, 95% CI: 2.0-4.2; *P* < 0.001). Among patients who were not prescribed statins prior to admission, those with HbA1c levels ≥ 6 exhibited the highest susceptibility to CHD, as indicated by an OR of 34.4 (95% CI: 4.2 to 283.6). However, it is important to note that the OR results displayed significant variability due to the limited sample size of the CHD group, which consisted of only 14 cases (**Figure 5**). In addition, HbA1c levels ≥ 6 also be recognized as a significant risk factor of AP patients who were not prescribed statins prior to admission (RO = 1.6, 95% CI: 1.1-2.3; *P* = 0.019) and individuals with hypertension extreme risk (RO = 2.7, 95% CI: 1.9-4.1; *P* < 0.001; Figure 5).

**Figure 4.**
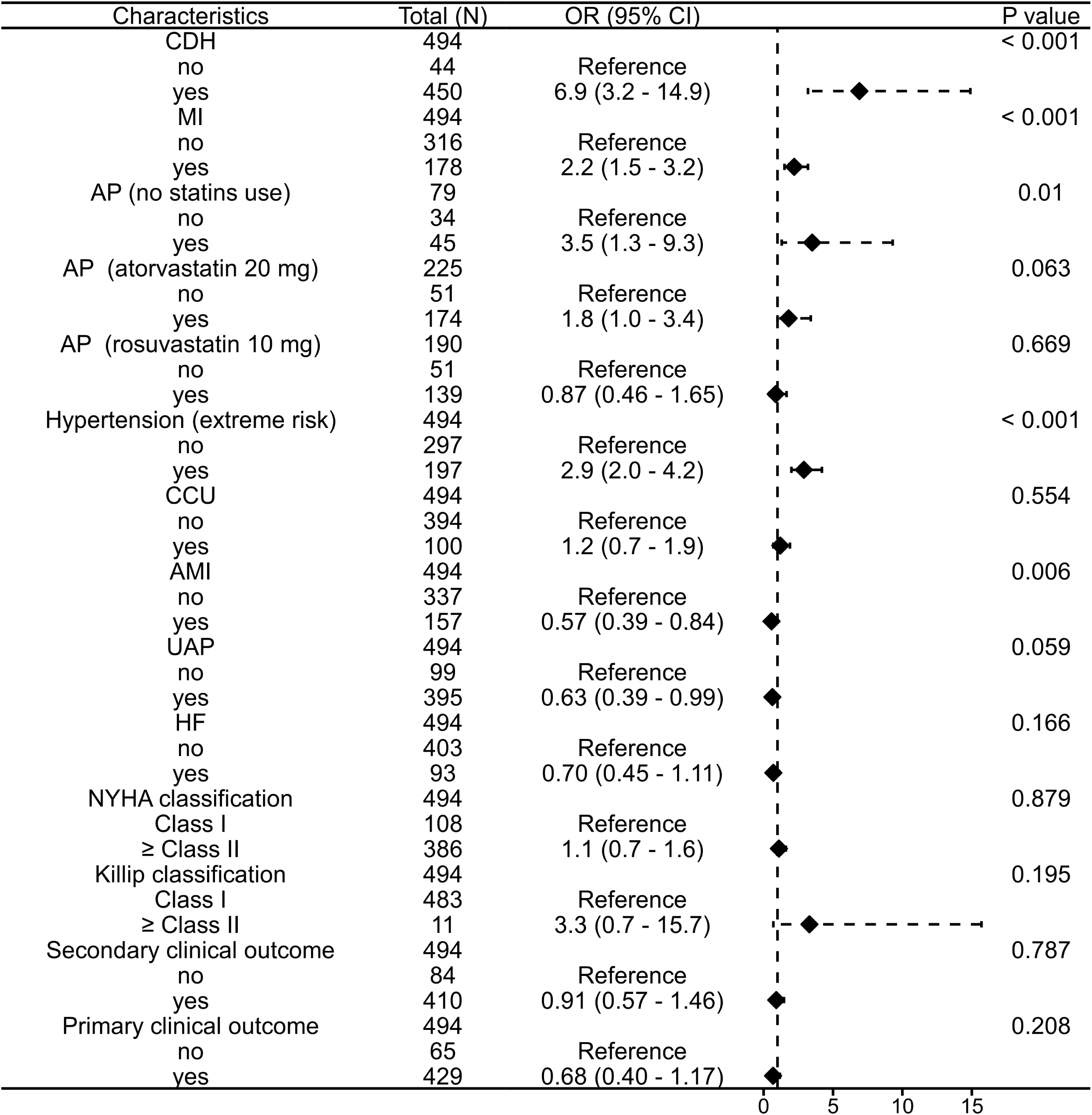
The impact of DM, stratified by statins use, on multiple clinical outcomes among CVD patients. OR, odds ratio; CDH, coronary heart disease; MI, myocardial infarction; AP, angina pectoris; DM, diabetes mellitus; CCU, coronary care unit; AMI, acute myocardial infarction; UAP, unstable angina pectoris; HF, heart failure.

**Figure 5.**
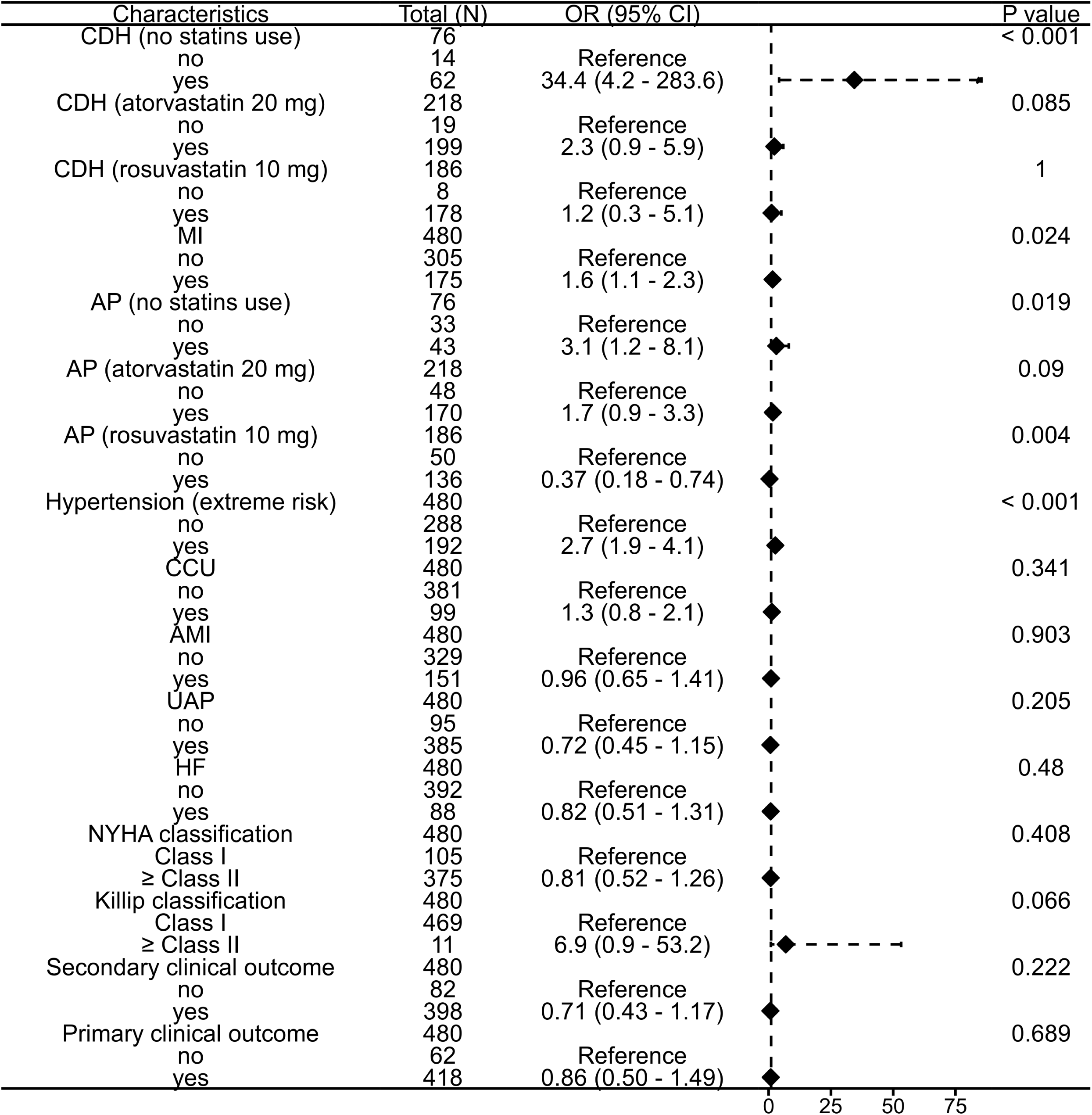
The impact of HbA1c (≥6% or < 6%), stratified by statins use, on multiple clinical outcomes among CVD patients. OR, odds ratio; CDH, coronary heart disease; MI, myocardial infarction; AP, angina pectoris; DM, diabetes mellitus; CCU, coronary care unit; AMI, acute myocardial infarction; UAP, unstable angina pectoris; HF, heart failure.

### Comparison of the effects of atorvastatin and rosuvastatin on multiple clinical outcomes in patients with CVD

The **Supplementary table 5** presents a comparison of the effects of atorvastatin and rosuvastatin on 13 clinical outcomes in patients diagnosed with CVD. In comparison to patients prescribed atorvastatin, those prescribed rosuvastatin exhibited the higher susceptibility to CHD and CCU, with OR values of 2.1 (95% CI: 1.0 to 4.2; *P* = 0.047) and 4.1 (95% CI: 2.5 to 6.6; *P* < 0.001), respectively. Additionally, the proportion of patients with NYHA classification and a lower risk of UAP were 0.51 (95% CI: 0.32 to 0.79; *P* = 0.003) and 0.62 (95% CI: 0.39 to 0.99; *P* = 0.043), respectively. In comparison to atorvastatin, the administration of rosuvastatin resulted in a statistically significant decrease in the median duration of hospitalization by one day, with a 95% CI ranging from 0.0 to 1.0 day (*P* = 0.001; **Supplementary table 6**).

### Interaction of statin use with hepatic FIB-4, complete blood count-derived inflammation indexes (CBCIIs) or ALBI grade

The baseline characteristics and statistical comparisons of both quantitative (**Supplementary table 7**) and qualitative (**Supplementary table 8**) parameters of individuals with CVD were using or not using statins. Significant differences were observed in height, weight, platelet count, ALT levels, and AST levels among the three groups (all P < 0.05). The use of statins led to a significant decrease in levels of TG, LDL-C, ApoB, and non-HDL-C (all P = 0.000) (**Supplementary table 7**). Analysis of the data in **Supplementary table 8** reveals statistically significant differences in gender, diabetes, and smoking history among the three groups (all P < 0.05). In order to assess the potential impact of statin use on improving clinical outcomes related to CVD by inhibiting the progression of liver fibrosis, we conducted a study to examine the association of different statin drugs and doses with FIB-4 levels in patients with CVD. The findings of our study demonstrated that atorvastatin was associated with lower FIB-4 levels in comparison to non-statin users, whereas rosuvastatin did not exhibit a significant difference in FIB-4 levels compared to non-statin users (**Supplementary table 9**). Our study also revealed that the use of statins was associated with a reduction in the levels of SIRI and SIRI/HDL-C, while also leading to an increase in PMR and SII in patients with CVD (**Supplementary table 9**). The findings of dose-stratification analysis in CVD patients revealed a correlation between the increased dosage of atorvastatin and rosuvastatin and the reduction in FIB-4 levels, suggesting the presence of a dose-response relationship (**Supplementary table 10**). Concurrently, a FIB-4 stratification analysis was conducted, revealing that patients treated with atorvastatin exhibited a higher proportion of FIB-4 < 1.45 and a lower proportion of FIB-4 > 3.25. Nevertheless, the reduction in FIB-4 level with rosuvastatin was not as pronounced as with atorvastatin (**Supplementary table 11**).

The ALBI grade is a liver function assessment tool that utilizes measurements of albumin and bilirubin levels. The study examined the impact of statin medications on liver function markers and ALBI grade. Statin use was found to significantly decrease DBIL levels compared to non-statin users, while showing no significant effect on other liver function indices, including ALBI grade (**Supplementary table 12**). Stratified analysis based on ALBI grade revealed that none of the patients progressed to an ALBI grade ≥-1.39, indicative of the group with the poorest prognosis. Furthermore, statin drug use did not influence ALBI grade in the remaining two groups (**Supplementary table 13**).

### Effect of statin use on atherogenic indices

As shown in **Supplementary table 14**, In comparison to statin-free group, the group with statin use exhibited a significant decrease in levels of AI, CRI-I, CRI-II, and LCI, whereas adjusted AIP, AIP, and RLP-C did not show a statistically significant difference.

## DISCUSSION

In our study, the baseline data analysis revealed that individuals with both HBV and CVD exhibited notably reduced levels of TG and Lp(a) compared to those with CVD alone. This observation could potentially be attributed to the varying prevalence of diabetes and the utilization of statins among the patient population. Consequently, it is imperative to stratify the HBV cohort based on their statin and diabetes risk profiles in order to comprehensively assess the diverse clinical outcomes in patients with CVD. The utilization of statins has the capacity to diminish the prevalence of CHD and AP among individuals with CVD associated with HBV infection. Stratification analysis, considering the presence of DM, revealed a notably heightened susceptibility to MI, AMI, and HF in patients with HBV infection combined with CVD. These findings imply that CVD patients with HBV infection should judiciously employ statins and effectively manage blood glucose levels.

HBV infection has the potential to give rise to a range of intrahepatic and extrahepatic ailments, encompassing acute and chronic hepatitis, cirrhosis, hepatocellular carcinoma, metabolic syndrome, CVD, and renal injury ^39–41^. The pathogenesis of HBV infection is intricate and multifaceted, essentially manifesting as a multisystem disorder. The underestimation of the influence of HBV infection on atherosclerosis has been observed in academic research. Multiple studies have demonstrated that the inflammation triggered by HBV infection serves as a significant contributing factor to the development of atherosclerosis. Conversely, atherosclerosis also exerts subsequent effects on the severity of chronic HBV infection, thereby establishing a detrimental cycle ^17^. In patients concomitant HBV and MAFLD decreases the risk of ASCVD when compared with MAFLD alone, which may attribute to antiviral treatment ^41^. The findings of present studies ^17, 41^ and our research indicate that the implementation of either antiviral therapy or lipid-lowering statin therapy holds the potential to improve liver metabolism or liver injury, consequently reducing the susceptibility to CVD associated with HBV infection.

Our study revealed that HBV infection posed a significant risk for AMI in patients with CVD (OR = 2.24, 95% CI: 1.40-3.57; *P* = 0.001). However, our findings did not indicate a significant association between HBV infection and MI (OR = 1.08, 95%CI: 0.69-1.68; *P* = 0.815). Analysis of baseline data demonstrated that the proportion of AMI in patients with both HBV infection and CVD was 39.1% (106/271), which was significantly higher than the proportion in patients with CVD alone (23.1%) (52/225; *P* < 0.001). Currently, there is a lack of pertinent research indicating that HBV infection serves as a risk factor for AMI. Furthermore, the underlying physiological and pathological mechanisms by which HBV infection may contribute to an increased risk of AMI in patients with CVD remain unclear. It is plausible that our findings may be attributed to potential data bias, as we did not conduct a multivariate regression analysis. Moreover, a comprehensive epidemiological investigation with larger sample sizes is necessary to ascertain the risk of HBV infection in relation to AMI. According to a comprehensive survey of over 90,000 individuals, patients with HBV and DM was found to be associated with significantly reduced risks of MI (aHR = 0.49; 95% CI: 0.42 to 0.56), ischaemic stroke (aHR = 0.61; 95% CI: 0.56 to 0.67), HF (aHR = 0.50; 95% CI: 0.43 to 0.59), and all-cause mortality (aHR = 0.72; 95% CI: 0.70 to 0.75) when compared to DM alone ^14^, while the researchers did not provide a specific explanation. Hence, the association between enhanced liver metabolism through antiviral therapy in patients with HBV and its potential correlation with MACE, HF, and all-cause mortality remains uncertain. In our study, the findings of risk stratification based on DM indicate that HBV infection is associated with a decreased risk of CDH in patients with CVD (OR = 0.32, 95% CI: 0.15-0.68; *P* = 0.004). However, when HBV infection is combined with DM, it is associated with an increased risk of MI (OR = 2.12, 95% CI: 1.30 – 3.45; P = 0.002), AMI (OR = 5.37, 95% CI: 2.91 – 9.92; *P* < 0.001), and HF (OR = 2.58, 95% CI: 1.32 - 5.04; *P* = 0.005).

These findings indicate that the clinical consequences linked to HBV infection in individuals with DM may have both positive and negative implications, contingent upon various confounding variables including the patient’s baseline characteristics, antiviral therapy, diabetes management, and statin administration. Furthermore, a comprehensive analysis was conducted on a cohort of 496 patients diagnosed with CVD to examine the variations in clinical and biochemical markers between individuals with CVD and DM and those without DM. Additionally, the impact of HbA1c levels and DM on various clinical outcomes in patients with CVD was investigated. Moreover, the effects of atorvastatin and rosuvastatin on multiple clinical outcomes in individuals with CVD were also examined. The correlation between DM and the extent of CVD has been examined in prior populations ^42^. The investigation revealed that DM is linked to a greater occurrence of cardiogenic shock, more severe stenosis (≥ 70%) segments, and a higher frequency of complications ^43^. A recent study encompassed a sample of 23,643 individuals who underwent consecutive coronary CT angiography. In comparison to non-DM patients, those with diabetes exhibited a heightened prevalence of obstructive CAD, characterized by more pronounced stenosis in both the proximal and middle segments of the coronary arteries ^44^. HbA1c serves as a precise diagnostic marker for diabetes, enabling the assessment of average blood sugar levels over a span of 2-3 months, while remaining minimally influenced by acute illnesses. Moreover, it holds the potential to inform treatment decisions. Subjects with HbA1c levels ranging from 5.7% to 6.4% exhibited elevated levels of coronary artery calcium and carotid intima-media thickness compared to subjects with HbA1c levels below 5.7%, suggesting that an increase in HbA1c levels is linked to an augmented burden of coronary and peripheral atherosclerosis in non-diabetic individuals with normal fasting glucose ^45^. A significant correlation was observed between the decrease in HbA1c levels and the reduction in HR for MACE ^46^. The meta-regression analysis further demonstrated that the mitigation of cardiovascular events among adult individuals with DM can be partially attributed to a decrease in HbA1c levels ^47^. Patients diagnosed with T2DM who exhibit inadequate control of their HbA1c levels (ranging from 8% to 8.9% and equal to or greater than 9%) are found to have comparable or elevated mortality risks when compared to patients diagnosed with CVD ^48^. Consequently, it is imperative to prioritize achieving optimal HbA1c levels in order to effectively mitigate risks, prevent mortality, and manage complications associated with DM.

The results of our study indicate that elevated HbA1c levels and the presence of DM have a significant influence on various clinical outcomes among individuals diagnosed with CVD. Specifically, our findings demonstrate that individuals with DM and HbA1c levels ≥ 6% are at an increased risk for developing CHD, experiencing MI, AP, and hypertension. Furthermore, our study examined the impact of both atorvastatin and rosuvastatin on various clinical outcomes among individuals diagnosed with CVD. Notably, patients administered rosuvastatin exhibited the greatest susceptibility to CHD and admission to the CCU, accompanied by elevated rates of NYHA classification. Conversely, these individuals experienced a diminished likelihood of developing UAP.

The findings of our study suggest an association between statin use and a decreased risk of HBV infection, potentially due to the inhibitory effects of statins on HBV-DNA replication or clearance. It may be prudent to consider reducing or discontinuing statin therapy in patients with HBV infection who exhibit transaminase levels exceeding three times the upper limit of normal. Hence, it is important to acknowledge the potential for bias in this conclusion. Additionally, the administration of anti-HBV therapy to patients with HBV infection could potentially impact the outcome. The quantification of HBV infection in this study was based on HBsAg levels, and while statin use may reduce HBV-DNA levels until they become undetectable, it is unlikely to result in clearance of HBsAg. Multiple studies have documented the potential of statins to decrease the likelihood of advancing to cirrhosis and hepatocellular carcinoma in individuals with HBV infection ^49–54^. This is attributed to the inhibition or eradication of HBV, which is believed to impede the progression to cirrhosis and liver cancer. Klundert et al. ^54^ discovered that statins inhibit the replication of the hepatitis B virus and decrease the viability of hepatoma cells by disrupting Rho-GTPases. Our findings align with this consensus. Furthermore, the HBV viral load remains relatively constant in the absence of effective anti-HBV treatment, suggesting that factors other than statins have minimal impact. Therefore, it is plausible to infer that the proposition of statins inhibiting or eradicating HBV is likely valid.

To delve deeper into the potential mechanisms of statin therapy in CVD patients to improve clinical outcomes, we conducted additional research on the impact of statin therapy on hepatic function, atherosclerotic burden, and systemic inflammation. The importance of FIB-4 and ALBI grade as the non-invasive indicators for liver fibrosis in predicting CVD among patients with liver diseases or DM has been well documented ^55–59^. Schonmann et al. ^58^ found that individuals with advanced fibrosis (FIB-4 ≥ 2.67) had a significantly higher risk of cardiovascular disease (HR = 1.60, 95% CI: 1.27-2.01) compared to those with inconclusive fibrosis (HR = 1.15, 95% CI: 1.01-1.31). Chun et al. ^59^ found a significant difference in the cumulative incidence of CVD among groups stratified by FIB-4. The severity of liver fibrosis was found to independently predict the occurrence of CVD in patients with T2DM. Additionally, the study revealed that the utilization of statins may serve as a protective factor (HR = 0.603) among individuals with T2DM, thereby mitigating the likelihood of developing CVD ^59^. FIB-4 may have utility in the identification of individuals at elevated risk of CVD within the population of patients diagnosed with NAFLD. Conversely, the use of APRI for this purpose is not advised ^60^. Our findings indicate that FIB-4 may be more effective than ALBI in assessing liver function in patients undergoing atorvastatin therapy. This is primarily due to the ability of FIB-4 to accurately monitor patients with severe liver damage, which was notably reduced in those receiving atorvastatin treatment. Prior research has indicated that statins may impact the levels of AST and ALT in NAFLD patients, as well as suggested that FIB-4 as a non-invasive test has an ability to predict hepatic fibrosis ^61^.

More importantly, our study revealed that both Atorvastatin and Rosuvastatin exhibited a dose-dependent reduction in FIB-4 levels among patients with CVD. A notable increase in the number of patients with FIB-4 grade < 1.45 and a decrease in those with FIB-4 grade > 3.25 were observed, particularly atorvastatin administration. Simon and colleagues ^62^ found that the use of statins was linked to a decrease in incident cirrhosis in a dose-dependent manner. Mohanty et al. ^63^ discovered that statin use in patients with HCV is correlated with a reduction of over 40% in the risk of cirrhosis. The prevalence of cirrhosis was found to be 8% lower in the group receiving statin treatment compared to the statins-free group ^64^. Furthermore, the use of statins was correlated with a reduced advancement of liver fibrosis and occurrence of hepatocellular carcinoma in a substantial population of Veterans with HCV infection ^64^. The collective findings of these studies ^62–64^ and our own research indicate that statins exhibit the most pronounced antifibrotic effects.

CBCIIs is an important predictor of chronic non-infectious inflammatory response, which is associated with the development of various metabolic system disorders such as fibrosis, hypertension, hyperlipidemia, and hyperglycemia ^65–67^. SIRI and SII are the two important inflammation indicators based on peripheral blood cell count ^32, 68^. Prospective investigations have identified a correlation between elevated levels of white blood cells and their subtypes, such as neutrophils, monocytes, and lymphocytes, with an augmented susceptibility to coronary heart disease and stroke. Neutrophils are capable of releasing a plethora of inflammatory mediators, chemokines, and oxygen free radicals, thereby instigating endothelial cell damage, consequent tissue ischemia, and a pivotal role in the inflammatory response of atherosclerosis ^32,68,69^. The activation and differentiation of monocytes into lipid-laden macrophages is a critical step in the pathogenesis of atherosclerotic plaques ^32,68,69^. Consequently, novel systemic inflammatory markers, such as the SII and SIRI, which are derived from neutrophil counts and other parameters, are likely to be correlated with unfavorable cardiovascular outcomes. In our study, contrasting outcomes were observed between SIRI and SII in assessing the impact of statin use in CVD patients. Specifically, our findings demonstrated a decrease in SIRI levels and an increase in SII levels in patients with CVD following statin therapy. Li et al. ^70^ demonstrated that SIRI is a strong predictor of all-cause mortality in CAD patients with low residual inflammatory risk. Numerous studies have indicated that the SII may serve as a potential biomarker for the development of CVD, with elevated levels of SII being linked to an increased risk of CVD ^71–73^. SII and SIRI have been linked to cardiovascular disease, yet the impact of statin therapy on these indices remains inconclusive. Following statin therapy, SII levels exhibited a significant increase, whereas SIRI levels showed a significant decrease. Consequently, the influence of statin therapy on the systemic inflammatory response in CVD patients remains uncertain and warrants further validation in a larger cohort.

Utilizing initial data, our study determined that the utilization of statins resulted in a significant decrease in TC and LDL-C levels. Additionally, we conducted a thorough examination of the impact of statin use on atherogenic indices among individuals diagnosed with CVD. Higher values of novel atherogenic indices, including AIP, CRI-I, CRI-II, and LCI, were found to be significantly associated with an increased risk of coronary artery disease (CAD) when compared to lower values ^35^. Many investigations indicate that the AIP serves as a novel marker in the contemporary landscape of CVD, with elevated AIP emerging as an independent prognostic factor in patients with CAD ^74–77^. AI was independently associated with all-cause and CVD mortality among patients undergoing peritoneal dialysis ^37^. Following adjustments for potential confounders, the highest of AI value exhibited a notably increased hazard ratio for both all-cause mortality and CVD mortality when compared to the lowest AI value ^37^. Hence, the majority of atherogenicity indicators hold significant implications for the diagnosis and prognosis of CVD risk events. Currently, there is a lack of relevant literature examining the relationship between statin use and atherogenicity indicators. Our research indicates that the use of statins has a significant impact on reducing levels of AI, CRI-I, CRI-II, and LCI. These findings suggest that statins may have a positive effect on liver fibrosis by improving lipid metabolism. Additionally, atherogenic indices such as AI, CRI-I, CRI-II, and LCI could serve as valuable markers for assessing liver fibrosis in patients undergoing statin therapy for CVD.

It is important to acknowledge that our study has several limitations. Firstly, the recruitment of patients was limited to a single center, and the sample size was moderate. Secondly, there is a dearth of information pertaining to the duration of hyperlipidemia, hypertension, and diabetes. Consequently, when assessing the impact of diabetes and its complications, it is not possible to disregard the potential influence of unmeasured confounding factors. Thirdly, the measurement of HbA1c may be susceptible to interference from hemoglobin variants, hereditary hemoglobinopathies, thalassemia, and iron-deficiency anemia. However, HbA1c serves as a reliable indicator of prolonged blood glucose regulation and exhibits reduced inter-individual variability in contrast to impaired glucose tolerance and fasting plasma glucose. Moreover, it is crucial to acknowledge that our analysis solely encompasses a single measurement of HbA1c, thereby rendering the alterations in glycemic control parameters indeterminate. Furthermore, this investigation solely encompasses inpatient data, precluding the assessment of long-term follow-up. Consequently, it is advisable to employ more sophisticated methodologies, such as cohort studies, to further investigate this matter.

## CONCLUSION

Our study indicates that CVD patients who also have HBV infection may experience a reduction in MACEs when treated with statins. DM with an HBA1c level of ≥ 6 is identified as risk factor for the development of CHD, MI, AP, and hypertension. The implementation of statins has demonstrated the potential to reduce the incidence of CHD and AP in individuals with CVD. The analysis of stratification, taking into account the presence of DM, has demonstrated a significantly increased vulnerability to MI, AMI, and HF among individuals with HBV infection combined with CVD. Furthermore, our findings reveal that long-term glycemic control should be prioritized in this patient population. Our findings also indicate that FIB-4 could serve as a potential indicator for assessing liver function in individuals undergoing treatment with statins. Statin therapy resulted in reduction in TC and LDL-C levels, as well as improvement of atherogenicity markers such as AI, CRI-I, CRI-II, and LCI. The results of our study indicate that statins may potentially ameliorate liver fibrosis through enhancement of lipid metabolism. Furthermore, atherogenic indices including AI, CRI-I, CRI-II, and LCI may serve as useful indicators for evaluating liver fibrosis in individuals receiving statin treatment for CVD. The observed improvements in hepatic function, atherosclerotic burden, and systemic inflammation associated with statin therapy may contribute to the favorable outcomes among individuals with CVD.

## Data Availability

Prof. Wang received grants from the Natural Science Foundation of Shaanxi Province, China. The authors declare that they have no competing interests.

## Supporting information

**Table S1-Table S14.**

